# Neuroimaging Summary Scores Predict Trajectories of Psychotic-Like Experiences in Youth

**DOI:** 10.64898/2026.06.03.26354754

**Authors:** Rebecca Cooper, Rashmi Sahasrabudhe, David C Glahn, Maria Jalbrzikowski

## Abstract

**Background:** Persistent, distressing psychotic-like experiences (PLEs) are associated with neurobiological alterations and increased psychosis risk. We combined individual-level neuroimaging measures with effect sizes from large neuroimaging studies to create a summary score (‘Psychosis Neuroscore’) reflecting neuroanatomic liability for psychosis, and examined its ability to predict PLE trajectories in young adolescents.

**Methods:** Using latent growth mixture models, we estimated PLE trajectories from four annual visits of the Adolescent Brain Cognitive Development Study (N=9584, ages 9-10 at baseline). Using baseline T1-weighted and diffusion-weighted imaging data, we calculated Psychosis Neuroscores, as well as Neuroscores for two psychiatric disorders with late adolescent/adult onset (Major Depressive Disorder, Bipolar Disorder). We compared Psychosis Neuroscores to i) other psychiatric Neuroscores, ii) modifiable risk factors, and iii) established risk factors in predicting trajectory membership.

**Results:** We identified four trajectories of distressing PLEs: Persistent Elevated (N=1,968, 21%), Gradual Decreasing (N=3,424, 36%), Rapid Decreasing (N=1,593, 17%) and Low/No Distress (N=2,599, 27%). Adolescents with Persistent Elevated PLEs had significantly higher Multimodal (combined T1 and diffusion-weighted) and T1-weighted Psychosis Neuroscores than all other trajectories (Odds Ratios [ORs] 1.27-1.34,*p*_FDR_<.01). Bipolar Disorder Neuroscores showed a similar pattern (ORs 1.16-1.23,*p*_FDR_<.01). Psychosis Neuroscores showed comparable associations with established risk factors in predicting trajectory membership, but smaller associations than modifiable risk factors, including screen time, physical activity, and sleep disturbances.

**Conclusions:** Psychosis Neuroscores differentiate youth with persistent PLEs from those with decreasing, remitting or low PLEs, demonstrating their potential utility for early risk stratification. Integration with established risk factors may enhance psychosis risk prediction in youth.

## Introduction

Psychotic-like experiences (PLEs) are unusual thoughts or experiences, such as mild perceptual abnormalities and delusion-like beliefs, that occur in the absence of a psychotic disorder diagnosis and at a subclinical level relative to the general population (1). PLEs are experienced by approximately 21% of adolescents (2), and are associated with adverse psychiatric, educational and functional outcomes, including suicidality, low educational attainment, and transdiagnostic psychiatric disorder diagnoses (1,3). Whereas 4.5-6% of youth experiencing PLEs later develop a psychotic disorder (4–6), PLEs that are persistent and distressing appear to be a more specific risk factor for psychosis (5,6). Identifying youth with persistent and distressing PLE trajectories is a critical first step in early intervention and prevention of psychosis.

Using data-driven modeling approaches, we recently identified four developmental trajectories of distressing PLEs in the Adolescent Brain Cognitive Development (ABCD) Study, a large, population-based sample of young adolescents (7). These trajectories comprised individuals experiencing distressing PLEs that were 1) consistently low (27% of the sample), 2) gradually decreasing (36%), 3) rapidly decreasing (17%) or 4) severe and persistent (21%) from late childhood through early adolescence (9-13 years). Youth experiencing severe and persistent trajectories endorsed engaging in longer amounts of screen time and having more disturbed sleep in comparison to the three other groups. Established risk factors for PLEs - including maternal pregnancy complications, experiences of bullying or trauma, and poorer cognitive performance - also predicted trajectory membership (7). While MRI has been considered a strong candidate for objective, biological psychosis risk assessments (8), the field has not yet successfully deployed neuroimaging with the balance of precision and feasibility necessary for individual psychosis risk detection, diagnosis, and/or treatment stratification (9). In other fields of medicine, when biological measures are included as a risk factor with other measures, risk prediction is significantly improved (10,11). Thus, combining neuroimaging markers with other established risk factors may enhance our ability to identify young people at greatest risk for developing psychosis.

PLEs are associated with diverse and widespread neurobiological alterations, including reductions in surface area, white matter and gray matter volume (12–14). These alterations are consistent with the pattern of alterations observed in individuals with psychosis (15), youth at clinical high risk for psychosis (16), and unaffected relatives of those with psychosis (17). However, the small effect sizes and distributed pattern of these associations makes it challenging to develop effective neuroimaging-based markers of psychosis risk. One way to address this challenge is to incorporate the distributed patterns of brain structural alterations associated with psychosis into a single summary risk score. This is conceptually similar to calculating polygenic risk scores in genetics, where an individual’s risk-associated genetic variants across the genome are weighted by an effect size for a particular phenotype and summed into a single risk score reflecting individual risk for the given phenotype (18). The genetic weights are derived from large, independent genome-wide association studies (GWAS (18)) related to this phenotype. Leveraging this approach in neuroimaging, we recently developed a neuroimaging summary score for psychosis, the “Psychosis Neuroscore”, using effect sizes from large-scale neuroimaging studies, i.e. the Enhancing NeuroImaging Genetics through Meta-Analysis [ENIGMA] consortium (15,19). We demonstrated that elevated Psychosis Neuroscores improves discrimination between individuals with schizophrenia and healthy controls in an independent sample (20). Using similar neuroimaging summary score approaches, others have shown neuroimaging scores are associated with diagnosis of psychosis in adults (21,22), family history of psychosis in youth (23), and are elevated in youth experiencing persistent distressing PLEs (24). Given the widespread neurobiological alterations associated with PLEs, extending this work by leveraging multimodal neuroimaging approaches to construct neuroimaging summary scores may improve our ability to identify youth with elevated and distressing PLEs. In addition, adopting a data-driven approach to characterize non-linear PLE trajectories may improve individualized risk prediction over threshold-based grouping methods. Comparing Psychosis Neuroscores to Neuroscores for other psychiatric disorders will determine the specificity of the Psychosis Neuroscore in indexing psychosis risk. Finally, comparing the Psychosis Neuroscore to other known risk factors for PLEs will help to contextualize findings, and help determine the predictive value of the Neuroscore relative to established risk factors.

Here, we leveraged data from ENIGMA consortium neuroimaging studies to develop multimodal neuroimaging summary scores for psychosis. Using data-driven trajectories we identified in previous work (7), we evaluated the ability of the Psychosis Neuroscore to predict longitudinal PLE trajectories across early adolescence. We calculated Neuroscores using multiple imaging modalities (T1-weighted, diffusion-weighted, and a combined multimodal approach) and compared their associations with trajectory membership. We evaluated the specificity of the Psychosis Neuroscore by calculating Neuroscores for major depressive disorder (MDD) and bipolar disorder, two psychiatric disorders with a median age of onset in late adolescence and adulthood (25) that have published ENIGMA group difference effect size maps, and compared their ability to predict trajectory membership. We also compared the Psychosis Neuroscore to an alternative neuroimaging summary score, the Regional Vulnerability Index (RVI) (22), in predicting PLE trajectories. Finally, we compared the predictive utility of the Psychosis Neuroscore to modifiable and established PLE risk factors in relation to trajectory membership. We expected that Psychosis Neuroscores would differentiate youth with Persistent Elevated PLEs from all remaining trajectories, with Multimodal Neuroscores demonstrating greater effect sizes than individual T1-weighted or diffusion-weighted Neuroscores. As persistent trajectories of PLEs appear to be more specific risk factors for psychotic disorders than for other psychiatric disorders (3,6,26), we expected that youth with Persistent Elevated PLEs would show higher Psychosis Neuroscores than Neuroscores for other psychiatric disorders.

## Methods and Materials

An overview of the study procedure is detailed in Figure 1.

**Figure 1.**
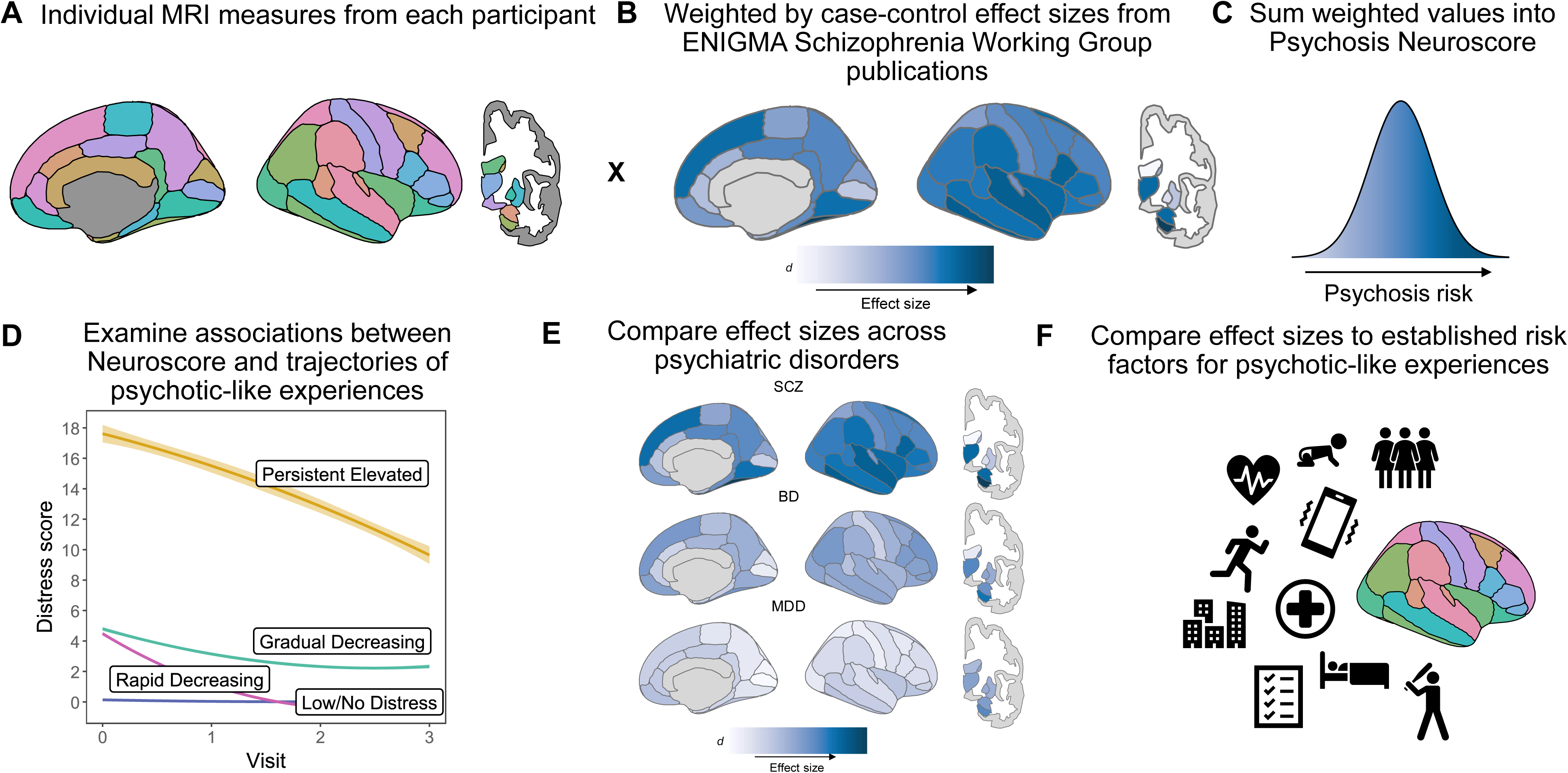
Study overview. **A.** Individual MRI measures, including surface area, cortical thickness, subcortical volume, and FA, were obtained for all participants. **B.** We multiplied individual MRI measures by Cohen’s *d* effect sizes for case-control differences in regional brain measures from the ENIGMA Schizophrenia working group. **C.** We summed the weighted values to create the Psychosis Neuroscore. We calculated separate Neuroscores using combinations of brain measures: T1-weighted (combination of surface area, cortical thickness, and subcortical volume), DTI (fractional anisotropy), and Multimodal (sum of T1-weighted and fractional anisotropy). We also calculated separate Neuroscores using individual brain measures (e.g., cortical thickness only). **D.** We used Multinomial logistic regressions to test associations between Psychosis Neuroscores and trajectories of distressing PLEs. **E.** We compared the Psychosis Neuroscore to Neuroscores for other psychiatric disorders in predicting trajectory membership. **F.** We compared the Psychosis Neuroscore to modifiable and established risk factors for PLEs in predicting trajectory membership. Abbreviations: BD, bipolar disorder; DTI, diffusion tensor imaging; MDD, major depressive disorder; MRI, magnetic resonance imaging; SCZ, schizophrenia.

### Participants

We used data from the Adolescent Brain Cognitive Development study (ABCD), a longitudinal cohort of 11,878 children recruited from 21 U.S. study sites assessed annually for up to 10 years (27). We included data from baseline, 1-year, 2-year and 3-year follow-up assessments (data release 5.1). As in our previous study (7), we excluded participants with missing data for the Prodromal Questionnaire-Brief Child Version (PQ-BC) at any assessment (N=1,790) and missing data for sex, site, race or ethnicity (N=3). From this sample (N = 10,075), which we used in our previous study (7), we further excluded individuals who did not pass quality control screening for T1-weighted neuroimaging data at baseline (N=493), resulting in a final analytical sample of N=9,584 (Table S1). When calculating the diffusion tensor imaging (DTI) and Multimodal Neuroscores, we also excluded participants that did not pass quality control screening for diffusion weighted neuroimaging data (N=2,003), resulting in N=7,581 participants in these analyses (Tables S2-S3). All individuals in the final analytical sample were also included in our previously published work (7). Institutional review boards approved study procedures at each site. Parents/guardians provided informed consent and children provided assent.

### Measures

#### Psychotic-Like Experiences

We used the PQ-BC to measure PLEs. The PQ-BC is a 21-item self-report questionnaire assessing psychotic-like phenomena and associated distress occurring in the previous 12 months (28). We calculated Distress scores according to previous approaches, summing the number of endorsed items and weighting by level of distress (range 0-6 from no distress to high distress; range 0-126). In our previous work we also calculated a Total score as the sum of all endorsed items; in this study, we focused on the Distress score as their trajectories are broadly similar and their respective associations with risk factors were nearly identical (7).

#### Image acquisition and processing

We used T1- and diffusion-weighted structural magnetic resonance images acquired on 3T scanners. T1-weighted data was processed following established protocols (detailed in Supplement). After excluding scans that did not pass quality control procedures (N=493), we extracted measures of surface area, cortical thickness and subcortical volume from the Desikan Killiany atlas for each image. We processed diffusion-weighted images using established pipelines (detailed in Supplement). After excluding scans that did not pass quality control measures (N=2,003), processed maps were inputted into Tract-Based Spatial Statistics (29), and projected onto the ENIGMA DTI skeleton. We extracted DTI measures from the overlap between the ENIGMA DTI skeleton and the JHU White Matter Label Atlas, resulting in estimates of fractional anisotropy (FA) for 17 bilateral tracts and three sections of the corpus callosum. Following our previous work and to minimize redundancy across DTI measures, DTI Neuroscores comprised only FA (20).

#### Neuroscore calculation

We calculated the Neuroscore by 1) regressing out the effects of age and sex for each brain region and z-scoring the residuals; 2) multiplying each residual by the corresponding regional Cohen’s *d* effect size (reflecting case-control differences) from ENIGMA Schizophrenia working group publications (15,19); and 3) summing the values across all brain measures to create a single Neuroscore for each individual, consistent with our previous work (20). Higher scores indicate a greater liability for psychosis. To calculate the T1-weighted Neuroscore, we used 152 regions of interest (68 surface area, 68 cortical thickness and 16 subcortical volume measures). See Figure S1 for a visualization of effect sizes for the T1-weighted Neuroscore. For the DTI Neuroscore, we used FA measures for 20 regions of interest. We also calculated separate Neuroscores for individual T1-weighted brain measures, including surface area, cortical thickness and subcortical volume. We then calculated a Multimodal Neuroscore by combining all T1-weighted and DTI measures (172 regions). Focusing on disorders with a median age of onset in late adolescence (25), we also used group difference effect sizes from the major depressive disorder (MDD) and bipolar disorder ENIGMA working group publications to calculate Multimodal, T1-weighted and DTI Neuroscores for these disorders (see Table S4).

#### Modifiable risk factors

We examined five modifiable risk factors - screen time, sleep disturbances, caffeine intake, lower physical activity and fewer recreational activities - in predicting PLE trajectories, which showed strong associations with PLE trajectories in our previous work (7). See Supplement for details on assessments used to measure these modifiable factors.

#### Established risk factors for PLEs

We identified ten risk factors for PLEs from previous work: family history of psychosis, obstetric and pregnancy complications, delayed developmental milestone achievement, traumatic experiences, migrant status, experiences of bullying, urban residence, and impairments in verbal memory and processing speed (30–33). We chose to focus on variables with <5% missingness, excluding delayed developmental milestones, urban residence, verbal memory and processing speed. See Supplement for details on assessments used to measure established PLE risk factors.

#### Regional Vulnerability Index

To provide additional context for our findings, we also calculated the Regional Vulnerability Index using T1-weighted measures. See Supplement for details on calculation.

### Statistical Analysis

Consistent with our previous work (7), we used latent growth mixture models to estimate PLE trajectories across four annual timepoints using Mplus (v8.11 (34)). We conducted a series of measurement invariance tests to evaluate whether the latent structure of the growth mixture model was statistically different from the structure identified in our previous paper (details in Supplement). Following model specification, each individual was assigned to a latent trajectory based on their highest class probability. We tested associations between trajectory membership and neuroimaging summary scores using multinomial logistic regressions. We ran these analyses using the bias-adjusted 3-step procedure (35), which accounts for uncertainty in class assignment and has been shown to reduce bias in parameter and standard error estimation compared to alternative approaches (35). This process involves 1) estimating an unconditional measurement model (i.e. the growth mixture model); 2) assigning individuals to their most likely latent class; and 3) conducting multinomial logistic regressions to assess the relationship between class assignment and predictor variables, taking into account the uncertainty in class assignment calculated in the second step. We ran separate models for each Neuroscore. Results are presented as Odds Ratios (ORs) with 95% confidence intervals (CIs), with the Low/No Distress trajectory as reference. We corrected for multiple comparisons (N=70) using the False Discovery Rate (36).

We compared the strength of associations between T1-weighted and DTI Psychosis Neuroscores and trajectory membership by including them simultaneously in a multivariable model predicting trajectory membership, represented by a latent variable. We tested equality of parameter coefficients using Wald tests, which evaluated the hypothesis *H*_0_: β_1_ - β_2_ = 0, where β_1_ and β_2_represent parameter coefficients for each predictor. Wald tests followed a ^2^ distribution with one degree of freedom. T1-weighted and DTI Neuroscores had variance inflation factors were lower than 1.5, indicating that multicollinearity was not evident when these measures were included jointly in the model. However, we did not compare any other pairs of Neuroscores as models showed strong evidence of multicollinearity (variance inflation factor >10). We also used Wald tests to compare Psychosis Neuroscores to several established risk factors and modifiable risk factors for PLEs identified in previous work (7). We ran separate multivariable models comparing Psychosis Neuroscores to a) established PLE risk factors and b) modifiable risk factors in predicting trajectory membership (as a latent variable). For these models, we compared all risk factors to the T1-weighted Psychosis Neuroscore to maximize sample size. To evaluate whether the Psychosis Neuroscore still predicted trajectory membership after adjustment for all other risk factors, we examined the effect sizes between Psychosis Neuroscores and trajectory membership while controlling for the effects of all other risk factors. We also used Wald tests to compare the Psychosis Neuroscore to the Regional Vulnerability Index in predicting trajectory membership. In sensitivity analyses, we i) additionally corrected for MRI scanner (rather than ABCD site), ii) additionally accounted for family relatedness by specifying clustering within families; and iii) examined more stringent statistical thresholds for including brain regions in Neuroscore construction.

## Results

The final sample comprised 9,584 participants (Table S1). T1-weighted Psychosis Neuroscores were weakly correlated with DTI Psychosis Neuroscores (*r*=0.14, *p*<2.2e-16). Multimodal Psychosis Neuroscores were highly correlated with the Multimodal Bipolar Disorder (*r*=0.92, *p*<2.2e-16). Post-hoc analyses examining individuals in the top 10% of Psychosis and Bipolar Disorder Neuroscores showed a substantial overlap of individuals identified in these two scores (72% overlap), indicating these scores largely captured the same individuals. Multimodal Psychosis Neuroscores were also correlated with MDD Neuroscores (*r*=0.60, *p*<2.2e-16).

### Characterization of PLE trajectories

Consistent with our previous work, we identified four trajectories that characterized a range of distressing PLEs: the first trajectory comprised youth experiencing few or no distressing PLEs (mean Distress score 0.15 [SD 0.35]), which we labeled as the ‘Low/No Distress’ trajectory (N=2,599, 27%); the second trajectory characterized youth experiencing mild-to-moderate distress associated with PLEs at baseline (mean Distress score 4.56 [SD 4.76]) that rapidly decreased throughout the study (labeled ‘Rapid Decreasing’; N=1,593, 17%); the third trajectory described youth experiencing mild-to-moderate distress at baseline (mean Distress score 4.83 [SD 6.09]), which decreased gradually over time (labeled ‘Gradual Decreasing’, N=3,424, 36%), and the fourth trajectory included youth that experienced elevated and persistent distressing PLEs (‘Persistent Elevated’, mean Distress score 17.38 [SD 16.19]; N=1,968, 21%; trajectories shown in Figure S2, descriptive statistics in Table S5). The number, variance, size and conceptual meaning of these trajectories was not statistically different from the trajectories identified in our previous paper (Supplemental Results & Table S6).

### Neuroimaging summary scores of psychosis risk predict persistent PLE trajectories

Individuals in the Persistent Elevated Distress trajectory had higher Multimodal and T1-weighted Neuroscores than individuals in all remaining trajectories (OR range 1.27-1.34, *p*_FDR_<.001; Figure 2A, Table 1). These individuals also had higher DTI Neuroscores than those in the Rapid Decreasing and Low/No Distress trajectories (ORs 1.15-1.18, *p*_FDR_≤.02). For individual measures contributing to the T1-weighted Neuroscore, all three measures - surface area, cortical thickness and subcortical volume - were significantly higher in the Persistent Elevated Distress trajectory compared to all remaining trajectories (ORs 1.10-1.46; Table S7, Figure S3). T1-weighted and DTI Neuroscores still significantly predicted trajectory membership even after mutually adjusting for each other in the same model (Figure S4).

**Figure 2.**
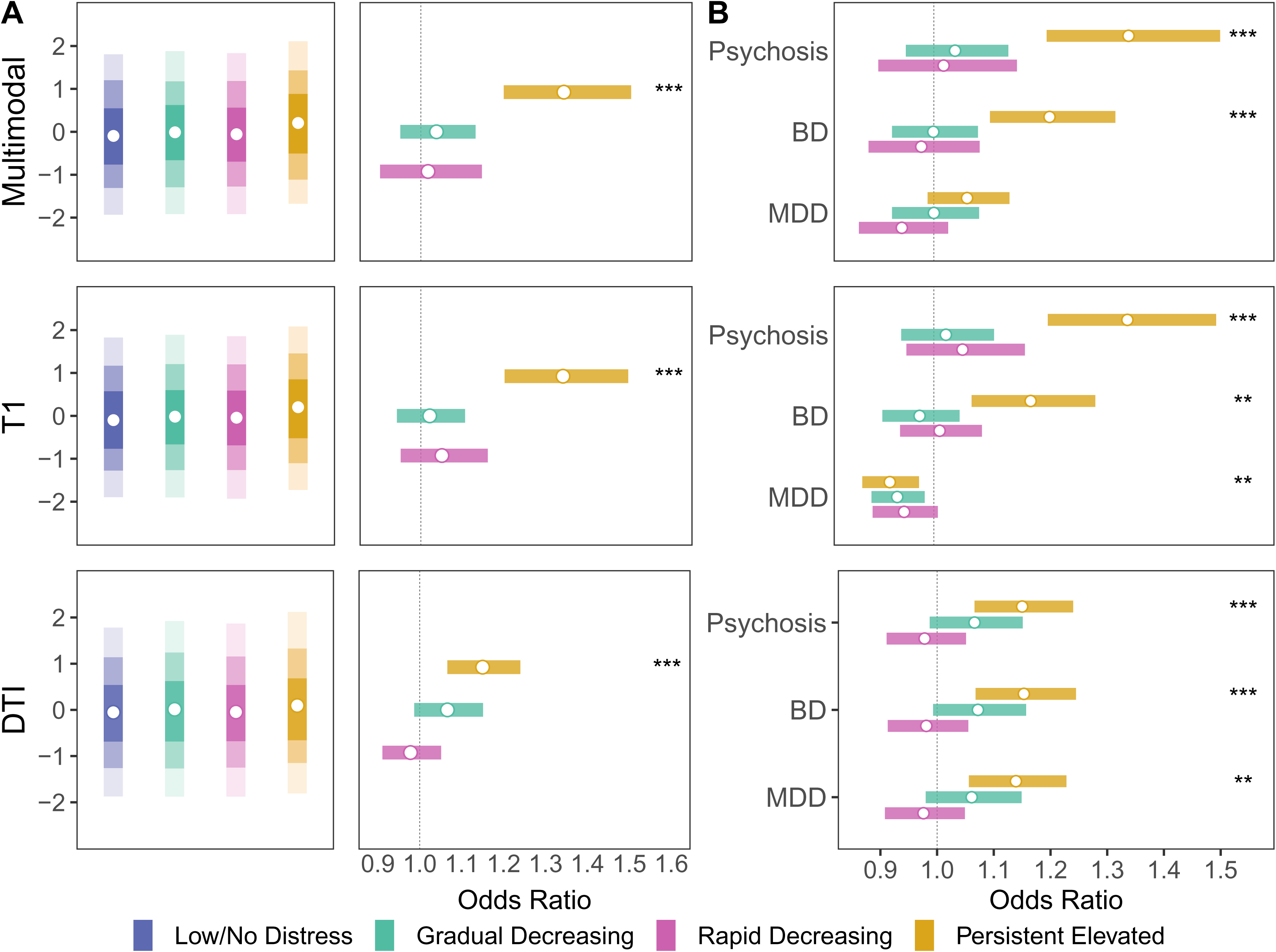
Associations between Neuroscores and trajectory membership. **A.** Left panels show distributions for Multimodal Psychosis Neuroscores (top), T1-weighted Psychosis Neuroscores (middle), and DTI Psychosis Neuroscores (bottom). Circles indicate mean values, shading indicates 50th (darkest) to 95th percentile of the distribution (lightest). Right panels show odds ratios for associations between each Distress trajectory and Psychosis Neuroscores, with the Low/No Distress trajectory as reference category. White circles indicate odds ratios; shaded bars indicate 95% confidence intervals. *=*p_FDR_*<0.05; **=*p_FDR_*<0.01, ***=*p_FDR_*<0.001. **B.** Panels show odds ratios for associations between each Distress trajectory and Multimodal Neuroscores (top), T1-weighted Neuroscores (middle), and DTI Neuroscores (bottom), for Psychosis Neuroscores, Bipolar Disorder Neuroscores, and MDD Neuroscores. Circles indicate mean values, shading indicates 50th (darkest) to 95th percentile of the distribution (lightest). The reference category is the Low/No Distress trajectory group. White circles indicate odds ratios; shaded bars indicate 95% confidence intervals. Abbreviations: BD, bipolar disorder; MDD, major depressive disorder.

**Table 1.**
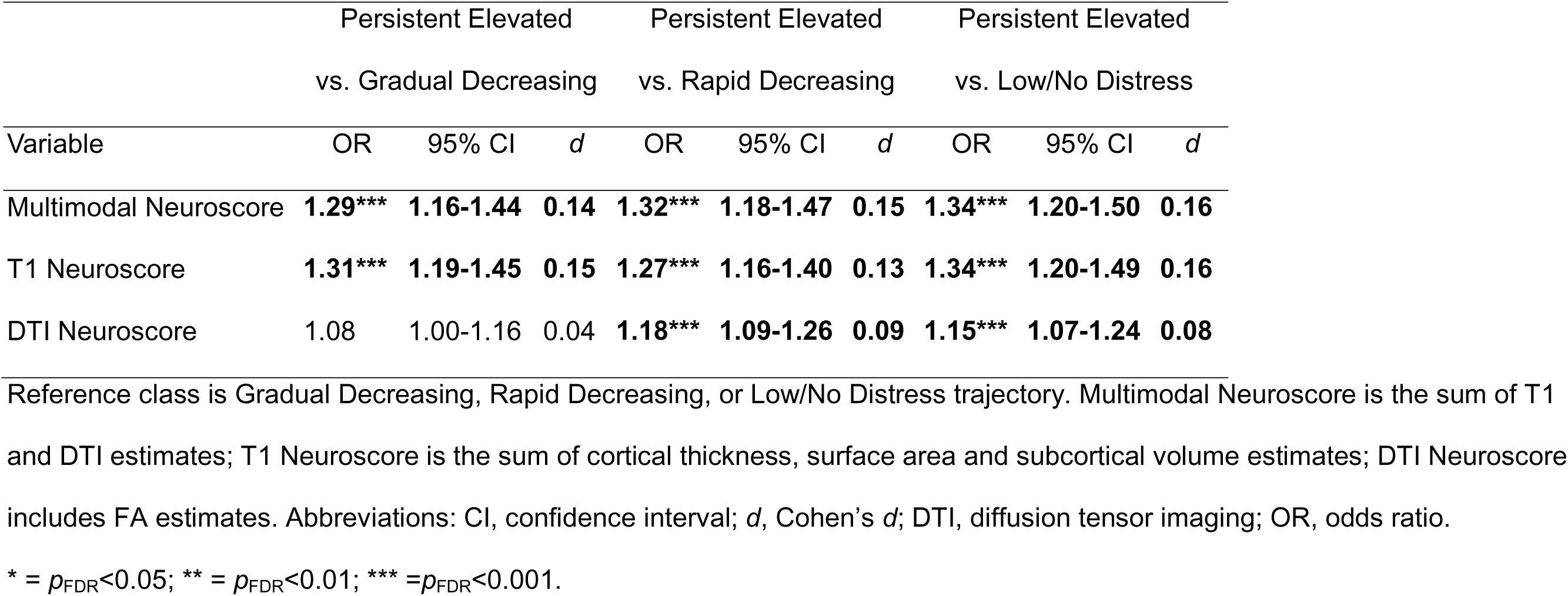
Associations between Psychosis Neuroscores and Distressing PLE trajectories.

|  | Persistent Elevated vs. Gradual Decreasing |  |  | Persistent Elevated vs. Rapid Decreasing |  |  | Persistent Elevated vs. Low/No Distress |  |  |
|---|---|---|---|---|---|---|---|---|---|
| Variable | OR | 95% CI | *d* | OR | 95% CI | *d* | OR | 95% CI | *d* |
| Multimodal Neuroscore | **1.29\*\*\*** | **1.16-1.44** | **0.14** | **1.32\*\*\*** | **1.18-1.47** | **0.15** | **1.34\*\*\*** | **1.20-1.50** | **0.16** |
| T1 Neuroscore | **1.31\*\*\*** | **1.19-1.45** | **0.15** | **1.27\*\*\*** | **1.16-1.40** | **0.13** | **1.34\*\*\*** | **1.20-1.49** | **0.16** |
| DTI Neuroscore | 1.08 | 1.00-1.16 | 0.04 | **1.18\*\*\*** | **1.09-1.26** | **0.09** | **1.15\*\*\*** | **1.07-1.24** | **0.08** |

Reference class is Gradual Decreasing, Rapid Decreasing, or Low/No Distress trajectory. Multimodal Neuroscore is the sum of T1 and DTI estimates; T1 Neuroscore is the sum of cortical thickness, surface area and subcortical volume estimates; DTI Neuroscore includes FA estimates. Abbreviations: CI, confidence interval; *d*, Cohen’s *d*; DTI, diffusion tensor imaging; OR, odds ratio.
\* = *p*<sub>FDR</sub><0.05; \*\* = *p*<sub>FDR</sub><0.01; \*\*\* =*p*<sub>FDR</sub><0.001.

### Psychiatric disorder neuroimaging summary scores predict longitudinal PLE trajectories

Individuals in the Persistent Elevated Distress trajectory had higher Multimodal, T1-weighted and DTI Bipolar Disorder Neuroscores (ORs 1.15-1.23) than those in all remaining trajectories (Figure 2B, Table 2). However, only select MDD Neuroscores distinguished the Persistent Elevated Distress trajectory from any of the other groups (Figure 2B, Table 2). Specifically, individuals in the Persistent Elevated Distress trajectory had higher Multimodal MDD and DTI MDD Neuroscores than the Rapid Decreasing trajectory (ORs 1.12-1.17), higher DTI MDD Neuroscores than the Low/No Distress and Rapid Decreasing trajectories (ORs 1.14-1.17) and lower T1-weighted MDD Neuroscores than the Low/No Distress trajectory (OR=0.92, *p*_FDR_=0.005).

**Table 2.** Associations between Psychosis, Major Depressive Disorder and Bipolar Disorder Neuroscores and Distressing PLE trajectories.

|  | Persistent Elevated<br>vs. Gradual Decreasing |  |  | Persistent Elevated<br>vs. Rapid Decreasing |  |  | Persistent Elevated<br>vs. Low/No Distress |  |  |
|---|---|---|---|---|---|---|---|---|---|
| Variable | OR | 95% CI | *d* | OR | 95% CI | *d* | OR | 95% CI | *d* |
| Multimodal Psychosis Neuroscore | **1.29\*\*\*** | **1.16-1.44** | **0.14** | **1.32\*\*\*** | **1.18-1.47** | **0.15** | **1.34\*\*\*** | **1.20-1.50** | **0.16** |
| T1 Psychosis Neuroscore | **1.31\*\*\*** | **1.19-1.45** | **0.15** | **1.27\*\*\*** | **1.16-1.40** | **0.13** | **1.34\*\*\*** | **1.20-1.49** | **0.16** |
| DTI Psychosis Neuroscore | 1.08 | 1.00-1.16 | 0.04 | **1.18\*\*\*** | **1.09-1.26** | **0.09** | **1.15\*\*\*** | **1.07-1.24** | **0.08** |
| Multimodal BD Neuroscore | **1.20\*\*\*** | **1.11-1.31** | **0.10** | **1.23\*\*\*** | **1.11-1.35** | **0.11** | **1.20\*\*\*** | **1.10-1.32** | **0.10** |
| T1 BD Neuroscore | **1.20\*\*\*** | **1.11-1.29** | **0.10** | **1.16\*\*** | **1.06-1.26** | **0.08** | **1.17\*\*** | **1.07-1.28** | **0.09** |
| DTI BD Neuroscore | 1.07 | 1.00-1.16 | 0.04 | **1.18\*\*\*** | **1.09-1.26** | **0.09** | **1.15\*\*\*** | **1.07-1.25** | **0.08** |
| Multimodal MDD Neuroscore | 1.06 | 0.98-1.14 | 0.03 | **1.12\*\*** | **1.05-1.20** | **0.06** | 1.06 | 0.99-1.13 | 0.03 |
| T1 MDD Neuroscore | 0.99 | 0.93-1.04 | -0.01 | 0.97 | 0.91-1.04 | -0.01 | **0.92\*\*** | **0.88-0.97** | **-0.04** |
| DTI MDD Neuroscore | 1.07 | 0.99-1.16 | 0.04 | **1.17\*\*\*** | **1.09-1.25** | **0.09** | **1.14\*\*** | **1.06-1.23** | **0.07** |

Reference class is Gradual Decreasing, Rapid Decreasing, or Low/No Distress trajectory. Multimodal Neuroscore is the sum of T1 and FA estimates; T1 Neuroscore is the sum of cortical thickness, surface area and subcortical volume estimates; DTI Neuroscore comprises FA estimates. Abbreviations: CI, confidence interval; *d*, Cohen’s *d*; DTI, diffusion weighted imaging; FA, fractional anisotropy; OR, odds ratio.
\* = *p*<sub>FDR</sub><0.05; \*\* = *p*<sub>FDR</sub><0.01; \*\*\* =*p*<sub>FDR</sub><0.001.

### Psychosis Neuroscores show comparable predictive ability to established PLE risk factors

Wald tests showed that T1-weighted Neuroscores and DTI Neuroscores showed comparable associations with latent trajectory membership (Wald ^2^=1.52, *p*_FDR_=0.245, Table S8). T1-weighted Neuroscores were also comparable to obstetric complications, trauma, family history of psychosis and migrant status in predicting trajectory membership (obstetric complications, Wald ^2^=2.46, *p*_FDR_=0.143; trauma, Wald ^2^=1.93, *p*_FDR_=0.189; family history of psychosis, Wald ^2^=3.71, *p*_FDR_=0.073; migrant status, Wald ^2^=4.10, *p*_FDR_=0.062; odds ratios shown in Figure 3A). Experiences of bullying and maternal pregnancy complications showed stronger relationships with PLE trajectories than Psychosis Neuroscores (all *p*_FDR_≤2.25e-04). T1-weighted and DTI Psychosis Neuroscores were still significantly associated with trajectory membership after adjusting for established and modifiable PLE risk factors (Figure 3B).

**Figure 3.**
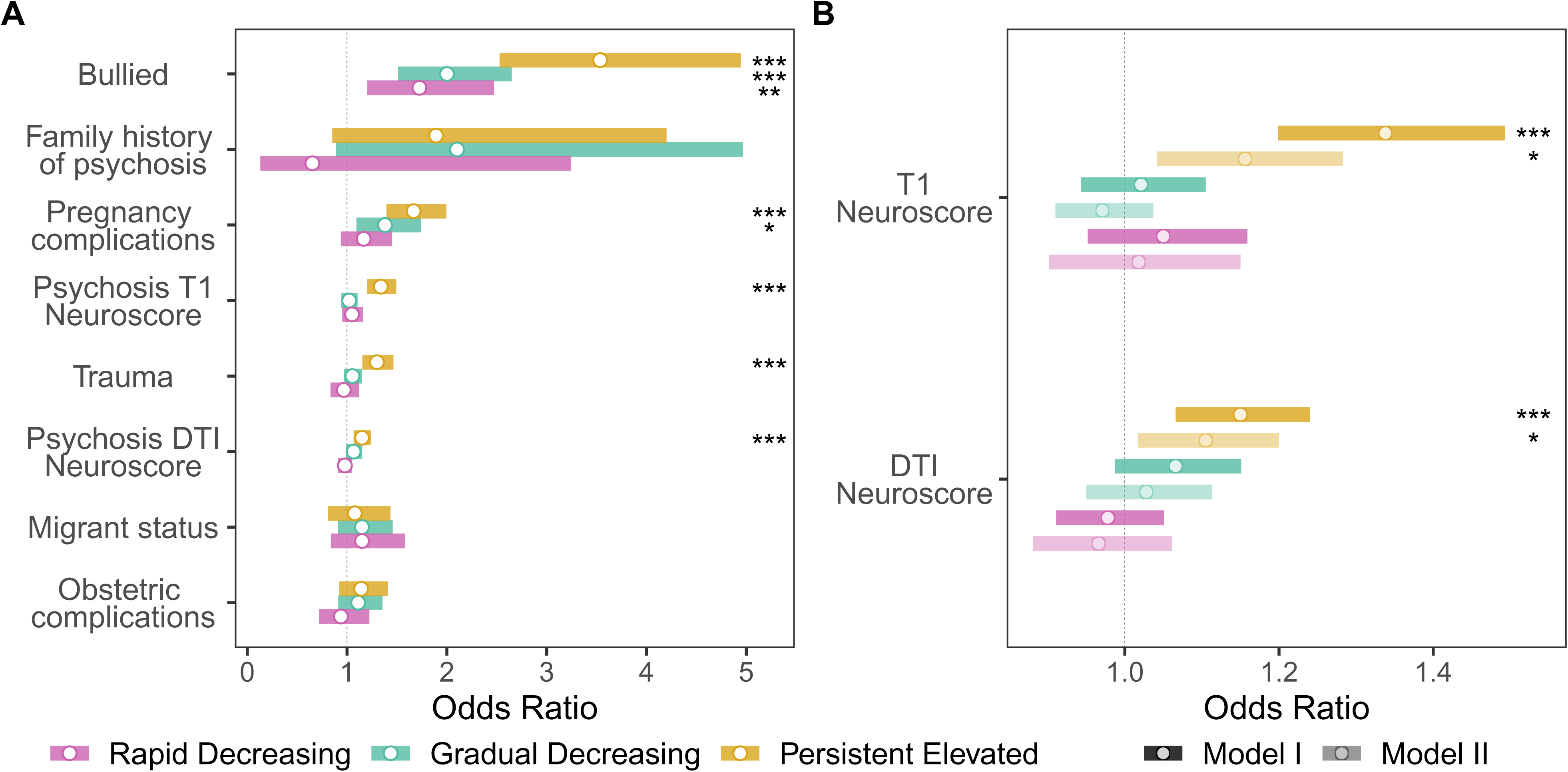
Associations between established risk factors and Psychosis Neuroscores and Distress PLE trajectory membership. **A.** Odds ratios for separate models examining associations between established risk factors, T1-weighted and DTI Psychosis Neuroscores, and Distress PLE trajectory membership. Circles represent odds ratios; bars represent 95% confidence intervals. Reference trajectory is the Low/No Distress trajectory**. B.** Odds Ratios for associations between T1-weighted and DTI Psychosis Neuroscores. Model I indicates original model; Model II adjusted for established risk factors (experiences of bullying, family history of psychosis, obstetric complications, pregnancy complications, migrant status, and traumatic experiences) and modifiable risk factors (sleep disturbances, caffeine intake, low physical activity, few recreational activities and screen time). Circles represent odds ratios; bars represent 95% confidence intervals. Reference trajectory is the Low/No Distress trajectory. * = *p*_FDR_<0.05; ** = *p*_FDR_<0.01; *** *p*_FDR_<0.001.

### Psychosis Neuroscores show comparable associations with select modifiable risk factors

T1-weighted Neuroscores were outperformed by four of five modifiable risk factors - screen time, sleep disturbances, lower physical activity and fewer recreational activities - in predicting trajectory membership (all *p*_FDR_≤0.010), but did not differ from caffeine intake (Wald ^2^=2.70, *p*_FDR_=0.126, Table S8; odds ratios and model coefficients shown in Figure S5).

### Psychosis Neuroscores outperform the Regional Vulnerability Index in predicting PLE trajectory membership

The Regional Vulnerability Index was moderately correlated with the T1-weighted Psychosis Neuroscore (*r*=0.62, p<2.2e-16) and was not associated with trajectory group membership (Table S9). Wald tests showed that the T1 Psychosis Neuroscore outperformed the Regional Vulnerability Index in predicting trajectory membership (Wald ^2^=5.30, *p*_FDR_=0.033).

Sensitivity analyses showed that results were highly consistent when we additionally accounted for family relatedness and MRI scanner (Tables S10-S11). Constructing Neuroscores using a reduced number of features (brain regions) by restricting the number of brain regions with increasingly more stringent statistical thresholds (top 75^th^, 50^th^ and 25^th^ percentile of brain regions) showed that reducing the number of features did not meaningfully influence our results (Table S12).

## Discussion

We found that Psychosis Neuroscores significantly differentiated youth with trajectories of persistent, distressing PLEs within a large, population-based adolescent cohort. Youth with severe and persistent distressing PLEs had Multimodal and T1-weighted Psychosis Neuroscores that were 13-33% higher than youth with decreasing, remitting or low PLE trajectories. Bipolar Disorder Neuroscores, but not MDD Neuroscores, also differentiated these youth from all remaining trajectories, suggesting Neuroscores show some specificity in reflecting psychiatric risk. Effect sizes for associations between Psychosis Neuroscores and PLE trajectory membership were comparable to some established risk factors for PLEs, including family history of psychosis, trauma, obstetric complications and migrant status. These findings extend previous work examining neuroimaging summary scores in adult samples, demonstrating their potential for improving risk prediction for severe psychiatric disorders in youth.

Psychosis Neuroscores were significantly higher in youth with persistent distressing PLEs than youth with decreasing, remitting or low levels of PLEs. Previously, the Psychosis Neuroscore has been used to distinguish adults with a psychotic disorder from healthy controls (20). In applying the Psychosis Neuroscore to a younger, general population sample, our findings indicate that the Neuroscore may not only help to differentiate case-control status but also capture groups considered to be high-risk for developing psychosis. The presence of elevated Neuroscores prior to psychosis suggests that the score may indicate mechanisms of risk. We look forward to future ABCD data releases to further assess the predictive performance of the Neuroscore in psychosis disorder onset as the ABCD participants reach late adolescence and early adulthood, the typical age of psychosis onset (25). Our findings align with prior work in the ABCD study showing that youth with persistent distressing PLEs show higher Regional Vulnerability Index scores than youth who did not experience PLEs (24). In the current study, however, the T1-weighted Regional Vulnerability Index did not predict trajectory membership (Table S9). The difference in findings may be explained by several methodological differences. First, the Regional Vulnerability Index and the Neuroscore measure distinct but related constructs: the Regional Vulnerability Index estimates the similarity between an individual’s brain structural measures and patterns evident in individuals with psychosis (22), whereas the Psychosis Neuroscore is a summed cross product of effect sizes, reflecting an individual’s putative neuroanatomical liability for psychosis. In addition, the previous study used a threshold-based approach to identify PLE groups, while we leveraged person-centered trajectory models, which enable us to characterize non-linear developmental trajectories and identify youth that may fall on the boundary of cut-off scores used in threshold-based approaches (37). These complementary approaches demonstrate the wealth of opportunities available for using diverse neuroimaging summary scores to identify youth at elevated risk for psychosis, and provide a foundation for future work seeking to develop objective predictors of psychosis risk in young adolescents.

Higher Neuroscores within the Persistent Elevated PLE trajectory indicate that subtle neuroanatomical alterations relevant to psychosis may be present in early adolescence, prior to psychotic disorder onset. These findings provide a launching point for further neuroanatomical investigations for psychosis risk in young people. In constructing Psychosis Neuroscores, both the pattern and overall distribution of effect sizes are important. Brain regions with larger schizophrenia case-control effect sizes are more heavily weighted, thus, there is a prominent role of frontal and temporal regions. Cortical thickness alterations in these regions are associated with conversion to psychosis in high-risk young people (16,38,39) and accelerated cortical thinning in frontal-temporal regions occurs prior to psychosis onset (40–42). Notably, cortical thickness changes in frontal and temporal regions also show the most protracted developmental trajectories across adolescence (43–45), highlighting the strong neurodevelopmental contribution of these regions to psychosis risk. Examining brain developmental changes across the PLE trajectories identified here, and how these changes may influence longitudinal changes in the Neuroscore, may reveal additional insight into neurodevelopmental mechanisms underpinning psychosis risk in young people.

Across modalities, we found that T1-weighted and DTI Psychosis Neuroscores were statistically equivalent in predicting trajectory membership, and were both significantly associated with trajectory membership even after mutually adjusting for one another in the same model. This suggests that the T1-weighted and DTI Neuroscores provide incremental, non-redundant information in predicting PLE trajectories. Nevertheless, the Multimodal Neuroscore provided little improvement in predicting PLE trajectories, showing similar odds ratios to the T1-weighted Neuroscore in differentiating trajectory groups. This suggests that incorporating diffusion-weighted data into Neuroscore calculations did not significantly improve prediction of PLE trajectories in this sample. Importantly, we note that the DTI subsample shows significant demographic differences from the full analytic sample. Thus, it is difficult to directly compare the performance of the Multimodal, DTI and T1 Neuroscores in this sample. An additional potential limitation of the DTI data is the use of the ENIGMA DTI processing pipeline, which is designed to facilitate streamlined analysis of large datasets but does not incorporate more computationally intensive quality control procedures, such as EPI susceptibility and B1 field inhomogeneity correction. Future studies should evaluate whether incorporating more advanced DTI preprocessing improves performance of the DTI Neuroscore.

Individuals with severe and persistent PLEs showed higher Bipolar Disorder, but not MDD Neuroscores than individuals in all remaining trajectories, indicating a degree of specificity in reflecting psychosis risk. Notably, this specificity was not previously observed in adults with schizophrenia, as Psychosis and MDD Neuroscores showed similar effect sizes in determining case-control status (20). This suggests Psychosis Neuroscores may have greater specificity at younger ages, although further investigation is required to confirm these findings. The similarity between Psychosis and Bipolar Disorder Neuroscores likely reflects their strong genetic, neurobiological, and clinical overlap (46,47). Indeed, multiple publications have identified strong relationships between the Bipolar Disorder and Schizophrenia working group effect size maps (*r*>0.70) (48,49). Prior work using alternative neuroimaging summary scores within the ABCD sample also showed that individuals with persistent distressing PLEs had higher summary scores for mood disorders, metabolic disorders, Parkinson’s and Alzheimer’s disease (24). Likewise, in adults, neuroimaging summary scores derived from functional network connectivity patterns associated with schizophrenia and bipolar disorder also show a high degree of overlap (50,51). Together, these findings indicate that brain structural correlates associated with psychiatric risk reflect a shared, underlying neurobiological basis (49). One way to capitalize on these findings would be to combine effect sizes across diagnostic categories into a single transdiagnostic Neuroscore, which would capture transdiagnostic risk across diverse psychiatric outcomes. Our group is actively investigating several methods to create transdiagnostic Neuroscores to improve early risk detection in young people.

At present, however, the substantial cost of acquiring neuroimaging data hinders accessibility and the potential to use the Neuroscore at scale for population-level risk prediction approaches. Furthermore, it may be more advantageous to incorporate Psychosis Neuroscores into sequential, multimodal workflows that incorporate neuroimaging data as needed, to further enhance the efficiency and decrease cost of early identification efforts (52). Alternatively, logistical and technological advances in accessible technologies, such as low-field MRI, have the potential to improve access to MRI in under-resourced settings (53,54). Our group found that low-field MRI shows strong correspondence (*r*>0.84) with research-grade MRI in estimating surface area and volume, demonstrating its potential for expansion beyond research settings (53). We are currently examining the validity of Neuroscores constructed from low-field MRI data as an important avenue for further research to integrate neuroimaging data into risk prediction models, ultimately improving early identification of severe mental illness.

Our findings contribute to the emerging literature using neuroimaging summary scores to improve prediction of psychiatric disorders (21,23,24,51,55–58). In the field of genetics, the performance of polygenic risk scores has been enhanced by using novel construction and modeling approaches, such as combining risk factor- and disease-specific polygenic risk scores (59), using statistical regularization techniques (60), incorporating functional annotations (47), and combining GWAS summary statistics across populations to enhance cross-ancestral accuracy (61). Translated to neuroimaging risk scores, this could involve combining structural and functional neuroimaging data (55), incorporating functional annotations (62), or using Bayesian methods to combine effect sizes (56,63). Thus, future work should focus on methodological enhancement of neuroimaging summary score performance, through additional fine-tuning and development.

The strongest predictors of trajectory membership were self-reported screen time, experiences of bullying, and maternal pregnancy complications, all of which significantly outperformed the T1-weighted Psychosis Neuroscore in predicting PLE trajectories. Screen time is associated with higher rates of social isolation, reduced sleep quality, and greater risk for cyberbullying victimization, which have been found to mediate associations between screen time and PLEs in young people (64,65). Social and cognitive abnormalities associated with PLEs may increase risk for bullying victimization (66). Pregnancy complications, such as maternal infections, may increase risk for abnormal neurodevelopmental trajectories associated with PLEs (32,67). However, T1-weighted Psychosis Neuroscores were still significantly associated with trajectory membership after inclusion of established risk factors, suggesting they may explain unique variance in predicting trajectory membership over and above established PLE risk factors. Taken together, these findings provide important context for interpreting the clinical relevance of Psychosis Neuroscores in risk prediction efforts. In future, combining Psychosis Neuroscores with established exposomic, genetic, clinical and demographic markers may further enhance psychosis risk prediction (68,69).

We note some limitations. PLEs were self-reported and not validated using clinical interviews, although the PQ-BC shows good correspondence with established psychosis symptom scales (e.g. the Structured Interview for Psychosis Risk Syndromes, SIPS) (70). Replication of these trajectories in an independent sample is an important future step to externally validate our findings. Extended longitudinal follow-up in the ABCD cohort will also be necessary to determine the long-term outcomes of each trajectory and the predictive role of the Neuroscore in predicting psychosis disorder onset. Participants excluded due to data missingness or during quality control processing were more likely to be of ethnic or racial minority and experienced more severe PLEs (Table S1), which could limit the generalizability of our findings, particularly when comparing the Neuroscore to established PLE risk factors. Novel quality control approaches could be used in future to retain youth not meeting conventional MRI quality control criteria (71).

In this study, we found that neuroimaging summary scores incorporating multimodal neuroimaging measures significantly differentiated youth with persistent, distressing PLEs from youth with decreasing, remitting or no distressing PLEs in a large, diverse cohort of young adolescents. Youth with persistent distressing PLEs also showed elevated T1-weighted and Multimodal Bipolar Disorder Neuroscores, but not MDD Neuroscores, reflecting greater specificity to psychosis. T1-weighted Psychosis Neuroscores showed comparable associations to specific established and modifiable risk factors for PLEs in predicting trajectory membership. Together, our findings suggest neuroimaging summary scores may help to identify youth at greatest risk for psychosis and could accelerate early identification and intervention efforts for serious mental illness in young people.

## Data Availability

All data from this study are available via the NIH Brain Development Cohorts Data Hub. Researchers can apply to use this data at https://www.nbdc-datahub.org/.

## Acknowledgements

RC was supported by the Tommy Fuss Center for Neuropsychiatric Disease Research Fellowship Award and the National Institutes of Mental Health (R01MH129636). MJ and DG were supported by the National Institutes of Mental Health (R01MH129636) and MJ was supported by the Tommy Fuss Center for Neuropsychiatric Research Next Generation Award. The study sponsors had no role in the study design; collection, analysis or interpretation of data; manuscript writing; or decision to submit the manuscript for publication.

We acknowledge the support of Boston Children’s Hospital’s High-Performance Computing Resources Clusters (Enkefalos 3) in conducting this research. Data used in the preparation of this article were obtained from the Adolescent Brain Cognitive Development^SM^ (ABCD) Study (https://abcdstudy.org), held in the NIMH Data Archive (NDA). This is a multisite, longitudinal study designed to recruit more than 10,000 children age 9-10 and follow them over 10 years into early adulthood. The ABCD Study® is supported by the National Institutes of Health and additional federal partners under award numbers U01DA041048, U01DA050989, U01DA051016, U01DA041022, U01DA051018, U01DA051037, U01DA050987, U01DA041174, U01DA041106, U01DA041117, U01DA041028, U01DA041134, U01DA050988, U01DA051039, U01DA041156, U01DA041025, U01DA041120, U01DA051038, U01DA041148, U01DA041093, U01DA041089, U24DA041123, U24DA041147. A full list of supporters is available at https://abcdstudy.org/federal-partners.html. A listing of participating sites and a complete listing of the study investigators can be found at https://abcdstudy.org/consortium_members/. ABCD consortium investigators designed and implemented the study and/or provided data but did not necessarily participate in the analysis or writing of this report. This manuscript reflects the views of the authors and may not reflect the opinions or views of the NIH or ABCD consortium investigators.

The ABCD data repository grows and changes over time. The ABCD data used in this report came from Data Release 5.1 (DOI <u>10.15154/z563-zd24</u>). The DOI for this study can be found at <u>10.15154/2q2s-vq77</u>.

CRediT author statement: **RC:** Methodology, Formal analysis, Investigation, Writing – Original Draft, Writing – Review & Editing, Visualization. **RS:** Formal Analysis, Writing – Review & Editing. **DG:** Writing – Review & Editing. **MJ:** Conceptualization, Methodology, Writing – Review & Editing, Supervision.

This manuscript was published as a preprint at https://doi.org/10.64898/2026.06.03.26354754.

