## Supplement for "Neuroimaging Summary Scores Predict Trajectories of Psychotic-Like Experiences in Youth"

Supplemental Methods.

**Measures**

*T1-weighted image acquisition and processing*

T1-weighted brain structural images were acquired on 3T magnetic resonance imaging (MRI) scanners across study sites and processed following unified protocols, as described previously (1,2). Briefly, images were processed using FreeSurfer (v7.1.1) (3). This involves removal of non-brain tissue, automated Talairach transformation, white matter segmentation, intensity normalization, tessellation of the gray/white matter boundary, automated topology correction, and surface deformation to delineate the gray/white matter and gray matter/cerebrospinal fluid boundaries. Images were then registered to a surface-based spherical atlas and parcellated using the Desikan Killiany atlas (4) into 34 regions per hemisphere (68 total). We obtained 68 cortical thickness measures and 68 surface area measures. We also extracted 16 subcortical volume measures from an atlas containing probabilistic information on the location of subcortical structures (i.e., aseg.mgz (5)). Quality control was conducted by ABCD investigators (detailed previously (2)). We used MRI scans that passed quality control criteria and were recommended for inclusion by the ABCD team. We then corrected for site and scanner differences using *Combat* (6), specifying age (non-linear effect) and sex (linear effect) as covariates.

*Diffusion data processing*

We processed diffusion-weighted data using the ENIGMA CHR DTI pipeline (7), which includes motion correction using FSL (8); eddy-current correction using FSL Eddy (8); diffusion tensor decomposition to derive fractional anisotropy (FA), axial diffusivity (AD), mean diffusivity (MD) and radial diffusivity (RD) maps. Using Psychiatry Neuroimaging Laboratory Tract-Based Spatial Statistics (PNL-TBSS) (9), diffusion tensor maps were projected onto the ENIGMA DTI skeleton, and DTI measures extracted from overlapping regions between the ENIGMA DTI skeleton and the JHU White Matter Atlas (10). We obtained fractional anisotropy estimates from 17 bilateral tracts and three sections of the corpus callosum, resulting in 20 DTI measures. We included MRI scans that passed quality control criteria that were recommended for inclusion by ABCD investigators. Following TBSS processing, we further excluded individuals with FA values >3SD from the mean (N=117). We then corrected for site and scanner differences using *Combat* (6), specifying age (non-linear effect) and sex (linear effect) as covariates.

*Modifiable risk factors*

We examined five modifiable risk factors that were associated with trajectory membership in our previous paper (11): screen time, sleep disturbances, caffeine intake, lower physical activity and fewer recreational activities.

Screen time was measured in the ABCD study using the 14-item self-report Screen Time Questionnaire (12). Participants reported the number of hours spent engaging with screen-based media on a typical weekday and weekend day (range 0-4+ hours). We combined these estimates to calculate a weighted average of screen time in a typical week.

We estimated sleep disturbances using the responses from a caregiver-reported 26-item Sleep Disturbance Scale for Children (13). We used the total score (sum of all items), with higher scores indicating greater sleep disturbance (range 26-126).

We used responses to the self-reported ABCD Substance Abuse questionnaire (14) to estimate weekly caffeine intake; participants reported the average number of caffeinated beverages they consumed per week over the past 6 months, which was multiplied by the standard caffeine content for each beverage to obtain an average weekly caffeine intake score.

We measured physical activity using responses to one self-report item from the Youth Risk Behavior Survey (15). Participants reported the number of hours in a typical week that they exercised for >1 hour. We reverse-scored the items so that higher scores indicate less physical activity.

We estimated recreational activities using responses from the caregiver-reported Sports and Activities Involvement Questionnaire (16). Caregivers reported on the number of recreational activities (such as sports and hobbies) their child had participated in within the previous 12 months. We reverse-scored this sum score so that higher scores indicate lower participation in recreational activities.

*Established risk factors for PLEs*

We examined the existing literature and identified ten established risk factors for PLEs: family history of psychosis, obstetric and pregnancy complications, delayed developmental milestone achievement, traumatic experiences, migrant status, experiences of bullying, urban residence, and impairments in verbal memory and processing speed (17–21). We focused on variables with <5% missingness, excluding delayed developmental milestones, urban residence, verbal memory and processing speed.

We assessed family history of psychosis using responses to the caregiver-reported ABCD Family History Assessment (16), which we scored as present if any first-degree relative experienced a psychotic disorder.

We used the caregiver-reported ABCD Developmental History Questionnaire (16) to assess obstetric and pregnancy complications. We coded as positive any endorsement of pregnancy complication (e.g., severe anemia or pregnancy-related diabetes) or obstetric complication (e.g., slow heartbeat at birth, or required oxygen at birth) and created a binary variable (yes=1, no=0).

We used the responses to the Traumatic Events module of the caregiver-reported Kiddie-Structured Assessment for Affective Disorders and Schizophrenia for DSM-5 (KSADS) (22) to assess traumatic experiences. This comprises 17 items assessing traumatic situations (e.g., a car accident, witnessed or caught in a fire). We scored all endorsed experiences as 1 and not endorsed as 0, and calculated a sum score representing the total number of traumatic experiences (possible range 0-17).

We assessed migrant status from the caregiver’s report of the child or parents’ country of birth. We coded migrant status as positive if the caregiver reported that the child or their parent was not born in the United States, creating a binary variable (yes=1, no=0).

To assess experiences of bullying, we used responses to one caregiver-reported item from the KSADS Background Items module (22). Caregivers reported whether their child experienced any problems with bullying at school or in their neighborhood (yes=1, no=0).

*Regional Vulnerability Index*

To calculate the Regional Vulnerability Index, we used the RVI_func() function from the R package *RVIpkg* (23). We used MRI data that were site- and scanner-corrected using Combat (6). Following methods described previously (24), we 1) regressed out the effects of age, sex, race, ethnicity and intracranial volume for each regional brain measure; 2) transformed the residuals using the inverse-normal transformation and standardized the transformed residuals; 3) calculated the Pearson correlation coefficient between the transformed residuals and the corresponding regional Cohen’s *d* effect size from ENIGMA Schizophrenia working group publications (25,26). Higher scores indicate greater neuroanatomical similarity to psychosis. We calculated an overall T1-weighted RVI by summing effect sizes for surface area, cortical thickness and subcortical volume values.

**Statistical Analysis**

*Growth mixture models*

We estimated trajectories of PQ-BC Distress scores using data from baseline and 1-, 2- and 3-year follow-up assessments. We modeled these trajectories using latent growth mixture models within Mplus (v8.11 (27)). Due to excess zeroes in Distress scores, we used zero-inflated negative binomial models to estimate these trajectories. Following recommended guidelines (28), we estimated models with one to six latent trajectory classes specifying either linear or quadratic growth parameters. We used both qualitative and quantitative criteria to select the best fitting model to the data: 1) Bayesian Information Criterion (BIC; lower values indicating better fit); 2) latent class size (all classes comprise >5% of the sample); 3) posterior probabilities (higher probabilities preferred); 4) Vuong-Lo-Mendell-Rubin and Lo-Mendell-Rubin likelihood ratio tests (p<.05 indicates current model is a better fit than a model with k-1 classes); and 5) model parsimony and interpretability.

*Model similarity tests*

Following recommendations (29), we estimated a series of sequential comparative tests to ensure the latent trajectory model identified in our previous study (N=10,075) was similar within the subsamples used in this study (T1 sample N=9,584; DTI sample N=7,581). We compared 1) the subsample used for T1 Neuroscore calculation (N=9,584) vs. individuals excluded from this sample but included in the previous paper (N=493), and 2) the subsample used for DTI Neuroscore calculation (N=7,581) vs. individuals excluded from this sample but included in the previous paper (N=2,494). We tested the assumptions of configural similarity (whether the optimal number of classes converge in each subsample), structural similarity (whether the profiles have similar meaning), dispersion similarity (whether the profiles have similar variance), and distributional similarity (whether the profiles are similar in size). To test configural similarity, we estimated a series of growth mixture models with one to six latent trajectory classes within each subgroup. We selected the optimal number of profiles using the same criteria that we used in our main analyses. We confirmed configural similarity if the subgroups converged on the same optimal number of profiles. Once confirmed, we tested for structural similarity by specifying each subgroup within a simultaneous multiple-group growth mixture model, and testing for equality of within-profile means. Once confirmed, we tested for dispersion similarity by testing for equality of within-profile variances. Finally, we tested distribution similarity by testing equality across class probabilities. For tests of structural, dispersion and distributional similarity, we used the Satorra-Bentler Scaled Χ^2^ test (30) to compare the fit of the nested model (parameters held equal) to the full model (parameters allowed to vary). A significant test statistic indicates the nested model has a significantly worse fit than the full model.

**Supplemental Results**

In testing configural similarity, the final analytic sample (N=9,584) and DTI subsample (N=7,581) showed optimal model fit when four Distress PLE trajectories were specified, consistent with the original sample from our previous paper (see Figure S1). This was indicated by clearly distinct trajectories and lower BIC. Tests for structural similarity, dispersion similarity and distributional similarity also indicated that the original analytic sample in our previous paper (N=10,055) and the final analytic sample in the current study (N=9,584) were statistically equivalent in i) within-profile means, ii) within-profile variances, and iii) relative sizes of trajectory groups (Table S4). This was also the case when comparing the final analytic sample (N=9,584) and the DTI subsample (N=7,581).

Table S1. Characteristics of included vs. excluded participants.

| N (%); Mean (SD) | Final analytic sample  N = 9,584^a^ | Excluded participants  N = 2,271^a^ | p-value^b^ |
| --- | --- | --- | --- |
| Sex assigned at birth |  |  | 0.6 |
| Female | 4,597 (48%) | 1,074 (47%) |  |
| Male | 4,987 (52%) | 1,197 (53%) |  |
| Age (baseline, months), Mean (SD) | 119.06 (7.52) | 118.65 (7.41) | 0.026 |
| Race, N (%) |  |  | <0.001 |
| White | 6,349 (66%) | 1,153 (51%) |  |
| Black/African American | 1,265 (13%) | 601 (26%) |  |
| American Indian/Alaskan | 45 (0.5%) | 17 (0.7%) |  |
| Pacific | 11 (0.1%) | 4 (0.2%) |  |
| Asian | 194 (2.0%) | 46 (2.0%) |  |
| Other/Multiple | 1,720 (18%) | 450 (20%) |  |
| Hispanic ethnicity, N (%) | 1,894 (20%) | 503 (23%) | 0.007 |
| Total PLE score, Mean (SD) | 2.56 (3.52) | 2.94 (3.72) | <0.001 |
| Distress PLE score, Mean (SD) | 6.06 (10.33) | 7.40 (11.66) | <0.001 |

^a^Final analytic sample vs. remainder of ABCD sample at baseline. ^b^Pearson's Chi-squared test (age, sex); Fisher's Exact Test (race); Wilcoxon rank sum test (continuous variables).

Table S2. Characteristics of DTI subsample vs. excluded participants.

| N (%); Mean (SD) | DTI subsample  N = 7,581^a^ | Excluded participants N = 4,274^a^ | p-value^b^ |
| --- | --- | --- | --- |
| Sex assigned at birth, N (%) |  |  | 0.2 |
| Female | 3,657 (48%) | 2,014 (47%) |  |
| Male | 3,924 (52%) | 2,260 (53%) |  |
| Age (baseline, months), Mean (SD) | 119.19 (7.56) | 118.59 (7.38) | <0.001 |
| Race, N (%) |  |  | <0.001 |
| White | 5,164 (68%) | 2,338 (55%) |  |
| Black/African American | 893 (12%) | 973 (23%) |  |
| American Indian/Alaskan | 31 (0.4%) | 31 (0.7%) |  |
| Pacific | 6 (<0.1%) | 9 (0.2%) |  |
| Asian | 156 (2.1%) | 84 (2.0%) |  |
| Other/Multiple | 1,331 (18%) | 839 (20%) |  |
| Hispanic ethnicity, N (%) | 1,473 (20%) | 924 (22%) | 0.003 |
| Total PLE score, Mean (SD) | 2.55 (3.52) | 2.77 (3.62) | <0.001 |
| Distress PLE score, Mean (SD) | 6.06 (10.43) | 6.78 (10.91) | <0.001 |

^a^DTI subsample vs. remainder of ABCD sample at baseline. ^b^Pearson's Chi-squared test (age, sex); Fisher's Exact Test (race); Wilcoxon rank sum test (continuous variables). Abbreviations: DTI, diffusion tensor imaging.

Table S3. Characteristics of DTI subsample vs. remaining participants.

|  | DTI subsample N = 7,581^a^ | Remaining participants N = 2,003^a^ | p-value^b^ |
| --- | --- | --- | --- |
| Sex assigned at birth, N (%) |  |  | 0.3 |
| Female | 3,657 (48%) | 940 (47%) |  |
| Male | 3,924 (52%) | 1,063 (53%) |  |
| Age (baseline, months), Mean (SD) | 119.19 (7.56) | 118.53 (7.35) | <0.001 |
| Race, N (%) |  |  | <0.001 |
| White | 5,164 (68%) | 1,185 (59%) |  |
| Black/African American | 893 (12%) | 372 (19%) |  |
| American Indian/Alaskan | 31 (0.4%) | 14 (0.7%) |  |
| Pacific | 6 (<0.1%) | 5 (0.2%) |  |
| Asian | 156 (2.1%) | 38 (1.9%) |  |
| Other/Multiple | 1,331 (18%) | 389 (19%) |  |
| Hispanic ethnicity, N (%) | 1,473 (20%) | 421 (21%) | 0.11 |
| Total PLE score, Mean (SD) | 2.55 (3.52) | 2.59 (3.50) | 0.5 |
| Distress PLE score, Mean (SD) | 6.06 (10.43) | 6.08 (9.95) | 0.3 |

^a^DTI subsample vs. remaining participants. All participants included in final analytic sample. ^b^Pearson's Chi-squared test (age, sex); Fisher's Exact Test (race); Wilcoxon rank sum test (continuous variables).

Table S4. Characteristics of studies used to create Psychiatric Neuroscores.

| Psychiatric disorder | Structures | N cases | N controls | Citation |
| --- | --- | --- | --- | --- |
| Psychosis | Cortical T1 | 4,474 | 5,098 | van Erp et al., 2018 (25) |
|  | Subcortical T1 | 2,028 | 2,540 | van Erp et al., 2016 (26) |
|  | DTI | 1,963 | 2,359 | Kelly et al., 2018 (31) |
| Major depressive disorder | Cortical T1 | 2,148 | 7,957 | Schmaal et al., 2017 (32) |
|  | Subcortical T1 | 1,728 | 7,199 | Schmaal et al., 2016 (33) |
|  | DTI | 1,265 | 921 | Van Velzen et al., 2020 (34) |
| Bipolar disorder | Cortical T1 | 1,837 | 2,582 | Hibar et al., 2018 (35) |
|  | Subcortical T1 | 1,710 | 2,594 | Hibar et al., 2016 (36) |
|  | DTI | 1,482 | 1,551 | Favre et al., 2019 (37) |

Abbreviations: DTI, diffusion tensor imaging.

Table S5. Characteristics of PLE trajectories.

| Characteristic | No Distress  N = 2,599 | Rapid Decreasing  N = 1,641 | Gradual Decreasing  N = 3,398 | Persistent Elevated  N = 1,946 |
| --- | --- | --- | --- | --- |
| Sex assigned at birth, N (%) |  |  |  |  |
| Female | 1,239 (48%) | 722 (44%) | 1,635 (48%) | 1,001 (51%) |
| Male | 1,360 (52%) | 919 (56%) | 1,763 (52%) | 945 (49%) |
| Missing | 0 | 0 | 0 | 0 |
| Age (baseline, months), Mean (SD) | 119.74 (7.57) | 119.31 (7.51) | 119.00 (7.51) | 118.02 (7.35) |
| Missing | 0 | 0 | 0 | 0 |
| Race, N (%) |  |  |  |  |
| White | 1,995 (77%) | 1,149 (70%) | 2,191 (64%) | 1,014 (52%) |
| Black/African American | 184 (7.1%) | 159 (9.7%) | 467 (14%) | 455 (23%) |
| American Indian/Alaskan | 7 (0.3%) | 5 (0.3%) | 16 (0.5%) | 17 (0.9%) |
| Pacific | 2 (<0.1%) | 4 (0.2%) | 3 (<0.1%) | 2 (0.1%) |
| Asian | 65 (2.5%) | 39 (2.4%) | 70 (2.1%) | 20 (1.0%) |
| Other/Multiple | 346 (13%) | 285 (17%) | 651 (19%) | 438 (23%) |
| Missing | 0 | 0 | 0 | 0 |
| Hispanic ethnicity, N (%) | 334 (13%) | 320 (20%) | 717 (21%) | 523 (27%) |
| Missing | 33 | 18 | 50 | 28 |
| Total PLE score, Mean (SD) | 0.15 (0.35) | 2.33 (2.09) | 2.42 (2.70) | 6.22 (4.81) |
| Missing | 0 | 0 | 0 | 0 |
| Distress PLE score, Mean (SD) | 0.15 (0.35) | 4.56 (4.76) | 4.83 (6.09) | 17.38 (16.19) |
| Missing | 0 | 0 | 0 | 0 |
| Sleep Disturbance total score, Mean (SD) | 35.18 (6.84) | 35.59 (7.40) | 36.51 (7.94) | 38.42 (9.75) |
| Missing | 3 | 6 | 6 | 6 |
| Physical activity (reverse-scored), Mean (SD) | 4.12 (2.25) | 4.34 (2.28) | 4.58 (2.29) | 4.83 (2.32) |
| Missing | 3 | 1 | 6 | 5 |
| Weekly caffeine intake, Mean (SD) | 1.26 (4.95) | 1.52 (4.34) | 1.86 (5.62) | 2.76 (7.77) |
| Missing | 0 | 0 | 3 | 2 |
| Weekly screen time, Mean (SD) | 2.80 (2.40) | 3.36 (2.75) | 3.75 (2.86) | 4.92 (3.60) |
| Missing | 3 | 1 | 3 | 3 |
| Recreational activities (reverse-scored), Mean (SD) | 19.14 (1.99) | 19.43 (2.02) | 19.61 (2.07) | 20.08 (1.86) |
| Missing | 0 | 0 | 0 | 0 |
| Trauma, N (%) | 0.40 (0.87) | 0.43 (0.82) | 0.48 (0.85) | 0.65 (0.99) |
| Missing | 65 | 49 | 92 | 61 |
| Family history of psychosis, N (%) | 29 (1.1%) | 22 (1.4%) | 79 (2.4%) | 66 (3.5%) |
| Missing | 63 | 47 | 88 | 58 |
| Pregnancy complications, N (%) | 926 (36%) | 662 (40%) | 1,496 (44%) | 952 (49%) |
| Missing | 0 | 0 | 0 | 0 |
| Obstetric complications, N (%) | 1,338 (51%) | 843 (51%) | 1,823 (54%) | 1,035 (53%) |
| Missing | 0 | 0 | 0 | 0 |
| Migrant status, N (%) | 401 (15%) | 295 (18%) | 662 (19%) | 407 (21%) |
| Missing | 0 | 2 | 3 | 3 |
| Bullied, N (%) | 221 (8.5%) | 228 (14%) | 544 (16%) | 436 (22%) |
| Missing | 1 | 0 | 0 | 1 |

Table S6. Model results for tests of configural, structural, dispersion, and distributional similarity across models including T1 (N=9,584) and DTI subsamples (N=7,581) vs. models that included individuals without neuroimaging data (N=10,075).

| Model | *np* | LL | AIC | BIC | Model test | ΔΧ^2^ | *df* | *p* |
| --- | --- | --- | --- | --- | --- | --- | --- | --- |
| *Final analytic sample* |  |  |  |  |  |  |  |  |
| M1: Configural | 45 | -86,647.89 | 173,385.80 | 173,710.60 |  |  |  |  |
| M2: Structural | 42 | -86,650.20 | 173,384.40 | 173,687.60 | M1 vs M2 | 3.57 | 4 | 0.567 |
| M3: Dispersion | 38 | -86,655.06 | 173,386.10 | 173,660.40 | M2 vs M3 | 3.45 | 5 | 0.631 |
| M4: Distributional | 35 | -86,656.65 | 173,383.30 | 173,635.90 | M3 vs M4 | 4.17 | 5 | 0.525 |
| *DTI subsample* |  |  |  |  |  |  |  |  |
| M1: Configural | 45 | -90,303.62 | 180,697.20 | 181,022.00 |  |  |  |  |
| M2: Structural | 42 | -90,305.24 | 180,694.50 | 180,997.60 | M1 vs M2 | 2.81 | 3 | 0.422 |
| M3: Dispersion | 38 | -90,311.07 | 180,698.10 | 180,972.40 | M2 vs M3 | 1.77 | 2 | 0.413 |
| M4: Distributional | 35 | -90,313.97 | 180,697.90 | 180,950.60 | M3 vs M4 | 4.53 | 4 | 0.339 |

Rows 1-5 show comparisons between original sample in previous study (N=10,055) and final analytic sample of current study (N=9,584). Rows 6-10 show comparisons between DTI subsample (N=7,581) and remaining individuals included within this study (N=2,003). The Satorra-Bentler Scaled Χ^2^ test was used to compare the fit of nested models(30). A non-significant result (p>.05) indicates that the more constrained (nested) model does not have a significantly worse fit than the less constrained (full) model. Abbreviations: AIC, Akaike Information Criterion; BIC, Bayesian Information Criterion, LL, Log-Likelihood; *np*, number of free parameters.

Table S7. Associations between individual measures contributing to T1-weighted Psychosis Neuroscores and Distressing PLE trajectories.

|  | Persistent Elevated vs. Gradual Decreasing | | | Persistent Elevated vs. Rapid Decreasing | | | Persistent Elevated  vs. Low/No Distress | | |
| --- | --- | --- | --- | --- | --- | --- | --- | --- | --- |
| Variable | OR | 95% CI | *d* | OR | 95% CI | *d* | OR | 95% CI | *d* |
| Cortical thickness Neuroscore | **1.14***** | **1.06-1.23** | **0.07** | **1.10*** | **1.02-1.19** | **0.05** | **1.10*** | **1.01-1.20** | **0.05** |
| Surface area Neuroscore | **1.33***** | **1.21-1.46** | **0.16** | **1.33***** | **1.21-1.48** | **0.16** | **1.46***** | **1.32-1.60** | **0.21** |
| Subcortical volume Neuroscore | **1.22***** | **1.11-1.34** | **0.11** | **1.23***** | **1.12-1.34** | **0.11** | **1.31***** | **1.19-1.44** | **0.15** |

Reference class is Gradual Decreasing, Rapid Decreasing, or Low/No Distress trajectory. Note that the cortical thickness, surface area and subcortical volume Neuroscores are summed to create the T1-weighted Psychosis Neuroscore. Abbreviations: CI, confidence interval; *d*, Cohen’s *d*; DTI, diffusion tensor imaging; OR, odds ratio.
* = *p*_FDR_<0.05; ** = *p*_FDR_<0.01; *** =*p*_FDR_<0.001.

Table S8. Results from Wald Χ^2^ tests testing equality of beta coefficients in predicting distressing PLE trajectory membership.

| Variable 1 | Variable 2 | Wald Χ^2^ test value | *df* | *p*_FDR_ |
| --- | --- | --- | --- | --- |
| *Neuroscores* |  |  |  |  |
| Psychosis T1 Neuroscore | Psychosis DTI Neuroscore | 1.52 | 1 | 0.245 |
| *Established risk factors* |  |  |  |  |
| Psychosis T1 Neuroscore | Bullied | 18.57 | 1 | **3.68e-05** |
| Psychosis T1 Neuroscore | Family history of psychosis | 3.71 | 1 | 0.073 |
| Psychosis T1 Neuroscore | Migrant status | 4.10 | 1 | 0.062 |
| Psychosis T1 Neuroscore | Obstetric complications | 2.46 | 1 | 0.143 |
| Psychosis T1 Neuroscore | Pregnancy complications | 15.06 | 1 | **2.25e-04** |
| Psychosis T1 Neuroscore | Trauma | 1.93 | 1 | 0.189 |
| *Modifiable risk factors* |  |  |  |  |
| Psychosis T1 Neuroscore | Caffeine intake | 2.70 | 1 | 0.126 |
| Psychosis T1 Neuroscore | Low physical activity | 7.13 | 1 | **0.012** |
| Psychosis T1 Neuroscore | Few recreational activities | 7.52 | 1 | **0.010** |
| Psychosis T1 Neuroscore | Screen time | 35.36 | 1 | **6.45e-09** |
| Psychosis T1 Neuroscore | Sleep disturbances | 10.86 | 1 | **1.80e-03** |

Risk factors were included in separate multivariable models predicting trajectory membership as a latent variable. Multivariable models were grouped into a) Neuroscores, b) established risk factors or c) modifiable risk factors. Within each model, pairs of variables (Variable 1 and Variable 2) were tested for parameter equality using Wald tests of the linear hypothesis *H*_0_: β_1_ - β_2_ = 0, which follows a Ꭓ^2^ distribution with one degree of freedom.

Table S9. Associations between T1-weighted Neuroscores, the Regional Vulnerability Index, and Distressing PLE trajectory membership.

|  | Persistent Elevated vs. Gradual Decreasing | | | Persistent Elevated vs. Rapid Decreasing | | | Persistent Elevated vs. Low/No Distress | | |
| --- | --- | --- | --- | --- | --- | --- | --- | --- | --- |
|  | OR | 95% CI | *d* | OR | 95% CI | *d* | OR | 95% CI | *d* |
| T1 Neuroscore | **1.31***** | **1.19-1.45** | **0.15** | **1.27***** | **1.16-1.40** | **0.13** | **1.34***** | **1.20-1.49** | **0.16** |
| Regional Vulnerability Index | 1.07 | 0.98-1.17 | 0.04 | 1.02 | 0.94-1.11 | 0.01 | 1.07 | 0.95-1.20 | 0.04 |

Reference class is Gradual Decreasing, Rapid Decreasing, or Low/No Distress trajectory. Abbreviations: CI, confidence interval; *d*, Cohen’s *d*; OR, odds ratio; T1, T1-weighted.
* = *p*_FDR_<0.05; ** = *p*_FDR_<0.01; *** =*p*_FDR_<0.001.

Table S10. Associations between Psychosis Neuroscores and Distressing PLE trajectories, clustering by family.

|  | Persistent Elevated vs. Gradual Decreasing | | | Persistent Elevated vs. Rapid Decreasing | | | | Persistent Elevated vs. Low/No Distress | | | |
| --- | --- | --- | --- | --- | --- | --- | --- | --- | --- | --- | --- |
|  | OR | 95% CI | *d* | | OR | 95% CI | *d* | | OR | 95% CI | *d* |
| Multimodal Neuroscore | **1.28***** | **1.16-1.42** | **0.14** | | **1.31***** | **1.17-1.47** | **0.15** | | **1.34***** | **1.21-1.48** | **0.16** |
| T1 Neuroscore | **1.30***** | **1.19-1.43** | **0.15** | | **1.27***** | **1.15-1.41** | **0.13** | | **1.34***** | **1.22-1.46** | **0.16** |
| Cortical thickness Neuroscore | **1.14*** | **1.04-1.25** | **0.07** | | **1.10*** | **1.00-1.22** | **0.05** | | **1.10*** | **1.01-1.20** | **0.05** |
| Surface area Neuroscore | **1.32***** | **1.20-1.45** | **0.15** | | **1.33***** | **1.20-1.48** | **0.16** | | **1.45***** | **1.33-1.59** | **0.21** |
| Subcortical volume Neuroscore | **1.20***** | **1.10-1.32** | **0.10** | | **1.23***** | **1.11-1.36** | **0.11** | | **1.30***** | **1.19-1.42** | **0.14** |
| DTI Neuroscore | 1.08 | 0.97-1.20 | 0.04 | | **1.18***** | **1.06-1.31** | **0.09** | | **1.14***** | **1.04-1.25** | **0.07** |

Reference class is Gradual Decreasing, Rapid Decreasing, or Low/No Distress trajectory. Multimodal Neuroscore is the sum of T1 and DTI estimates; T1 Neuroscore is the sum of cortical thickness, surface area and subcortical volume estimates; DTI Neuroscore includes fractional anisotropy estimates. Abbreviations: CI, confidence interval; *d*, Cohen’s *d*; DTI, diffusion tensor imaging; OR, odds ratio.
* = *p*_FDR_<0.05; ** = *p*_FDR_<0.01; *** =*p*_FDR_<0.001.

Table S11. Associations between Psychosis Neuroscores and Distressing PLE trajectories, correcting for scanner.

|  | Persistent Elevated vs. Gradual Decreasing | | | Persistent Elevated vs. Rapid Decreasing | | | | Persistent Elevated vs. Low/No Distress | | | |
| --- | --- | --- | --- | --- | --- | --- | --- | --- | --- | --- | --- |
|  | OR | 95% CI | *d* | | OR | 95% CI | *d* | | OR | 95% CI | *d* |
| Multimodal Neuroscore | **1.28***** | **1.16-1.41** | **0.14** | | **1.32***** | **1.18-1.48** | **0.15** | | **1.34***** | **1.21-1.48** | **0.16** |
| T1 Neuroscore | **1.30***** | **1.20-1.41** | **0.14** | | **1.28***** | **1.16-1.41** | **0.13** | | **1.33***** | **1.21-1.47** | **0.16** |
| Cortical thickness Neuroscore | **1.14***** | **1.07-1.21** | **0.07** | | **1.10*** | **1.02-1.20** | **0.06** | | **1.10*** | **1.01-1.20** | **0.05** |
| Surface area Neuroscore | **1.31***** | **1.20-1.43** | **0.15** | | **1.34***** | **1.20-1.49** | **0.16** | | **1.45***** | **1.33-1.58** | **0.20** |
| Subcortical volume Neuroscore | **1.20***** | **1.10-1.32** | **0.10** | | **1.23***** | **1.12-1.35** | **0.11** | | **1.30***** | **1.17-1.43** | **0.14** |
| DTI Neuroscore | 1.07 | 0.98-1.16 | 0.04 | | **1.20***** | **1.11-1.30** | **0.10** | | **1.14***** | **1.04-1.24** | **0.07** |

Reference class is Gradual Decreasing, Rapid Decreasing, or Low/No Distress trajectory. Abbreviations: CI, confidence interval; *d*, Cohen’s *d*; DTI, diffusion tensor imaging; OR, odds ratio.
* = *p*_FDR_<0.05; ** = *p*_FDR_<0.01; *** =*p*_FDR_<0.001.

Table S12. Associations between T1 Psychosis Neuroscores and Distressing PLE trajectories, using a reduced number of significant brain regions to construct the Neuroscore.

| P-value threshold^a^ | N regions | Persistent Elevated vs. Gradual Decreasing | | | Persistent Elevated vs. Rapid Decreasing | | | Persistent Elevated vs. Low/No Distress | | | |
| --- | --- | --- | --- | --- | --- | --- | --- | --- | --- | --- | --- |
|  |  | OR | 95% CI | *d* | OR | 95% CI | *d* | | OR | 95% CI | *d* |
| None | 152 | **1.31***** | **1.19-1.45** | **0.15** | **1.27***** | **1.16-1.40** | **0.13** | | **1.34***** | **1.20-1.49** | **0.16** |
| Top 75th percentile | 114 | **1.29***** | **1.17-1.41** | **0.14** | **1.24***** | **1.13-1.36** | **0.12** | | **1.30***** | **1.17-1.45** | **0.15** |
| Top 50th percentile | 76 | **1.21***** | **1.11-1.31** | **0.11** | **1.16***** | **1.08-1.26** | **0.08** | | **1.20***** | **1.08-1.32** | **0.10** |
| Top 25th percentile | 38 | **1.16***** | **1.08-1.25** | **0.08** | **1.13**** | **1.05-1.21** | **0.07** | | **1.14**** | **1.04-1.25** | **0.07** |

^a^T1 Psychosis Neuroscores were constructed using subsets of brain regions according to the statistical significance of their corresponding effect sizes in the ENIGMA Schizophrenia Working Group publications. Only the top (75^th^, 50^th^ or 25^th^) percentile of regions that differentiated between cases and controls were used to construct the Neuroscore. Reference class is Gradual Decreasing, Rapid Decreasing, or Low/No Distress trajectory. Abbreviations: CI, confidence interval; *d*, Cohen’s *d*; OR, odds ratio.
* = *p*_FDR_<0.05; ** = *p*_FDR_<0.01; *** =*p*_FDR_<0.001.

Figure S1. Effect sizes used to calculate the T1-weighted Psychosis Neuroscore.

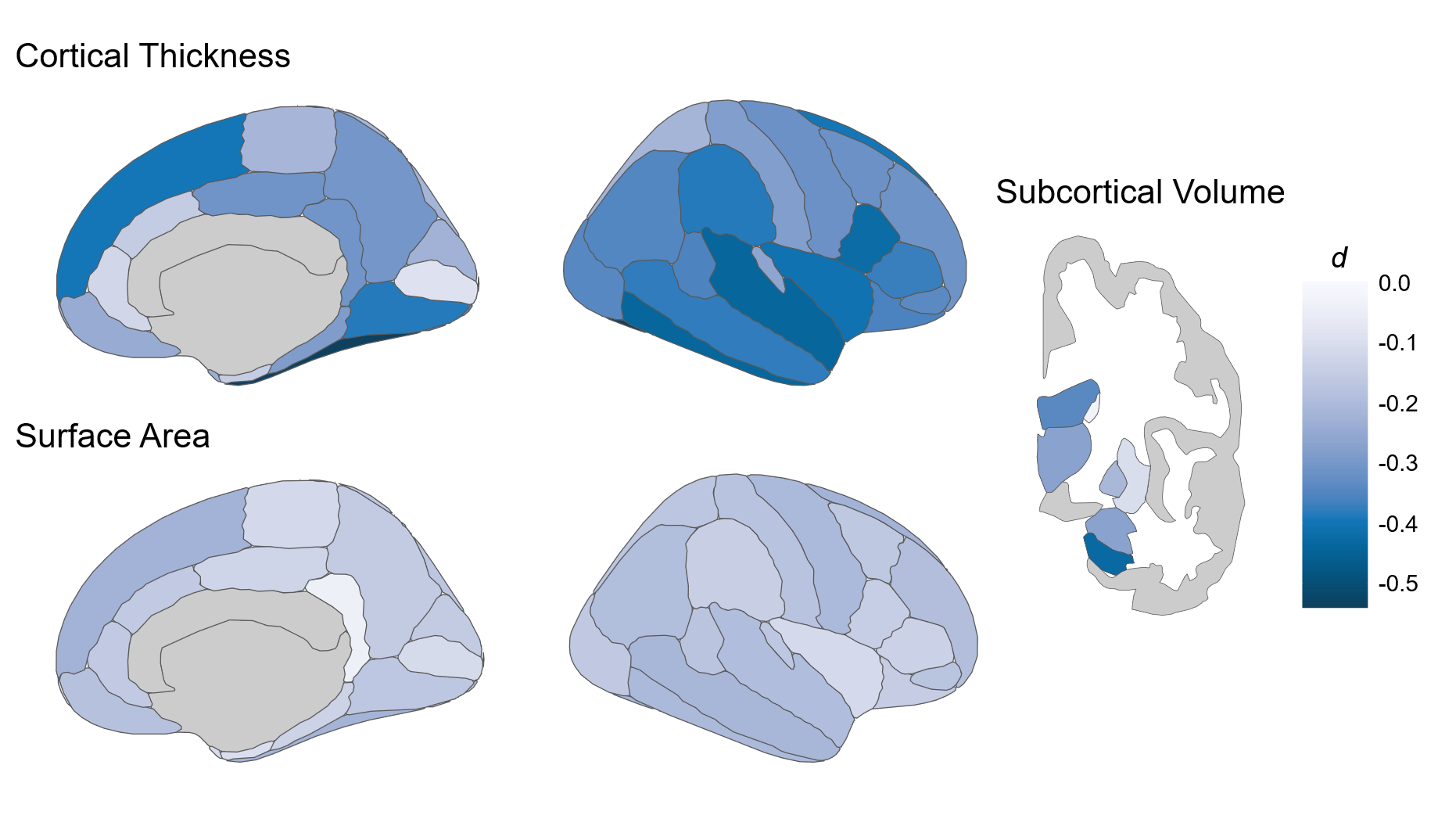

Effect sizes obtained from ENIGMA Schizophrenia Working Group publications (25,26). Darker color indicates larger group difference effect sizes between schizophrenia cases and healthy controls. Abbreviations: *d*, Cohen’s *d* effect size.

Figure S2. Comparison of PLE trajectories in the previous study and in the final analytic sample.

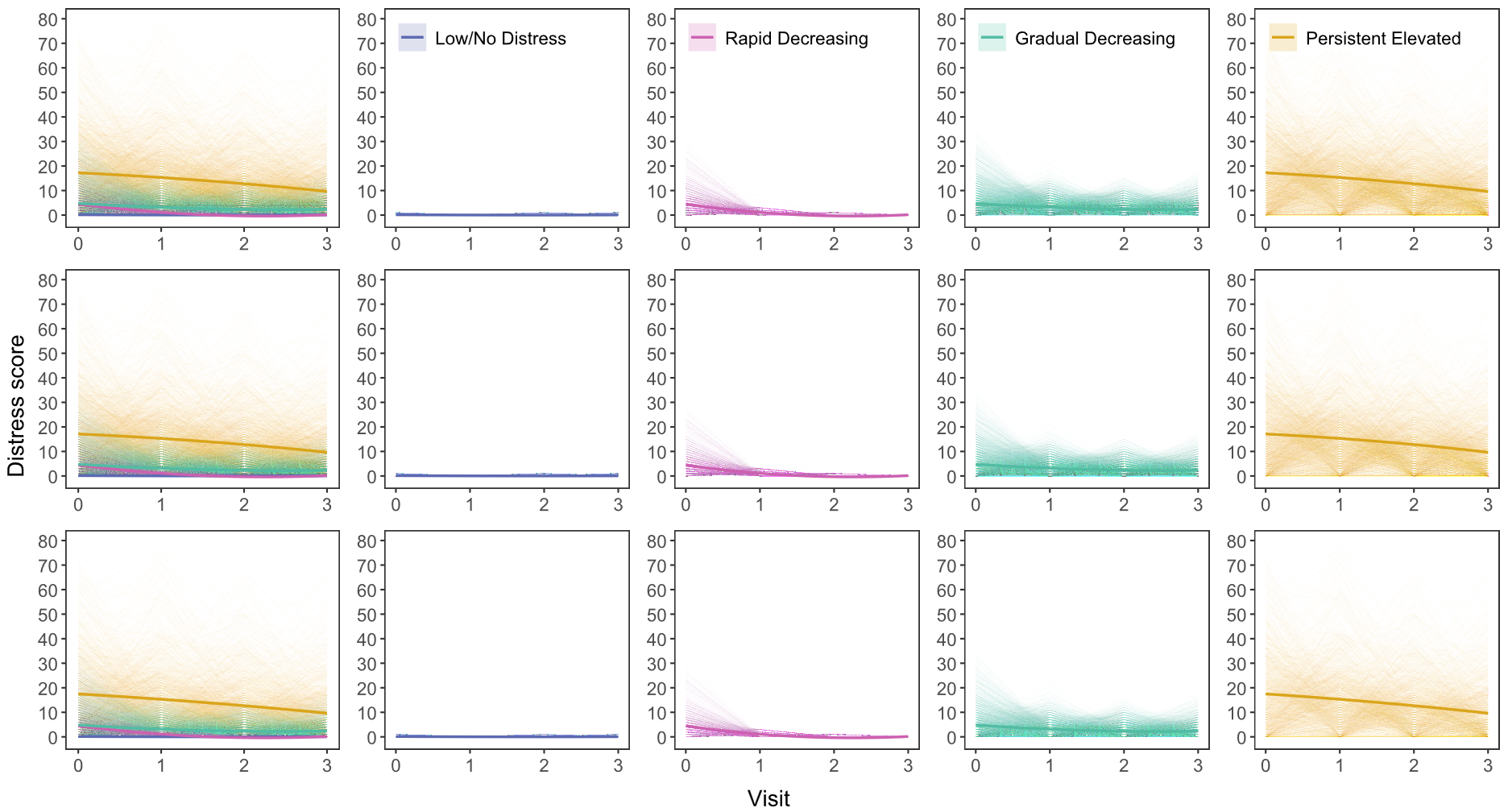

First row: Distress trajectories identified in our previous study(11). Second row: Distress trajectories within the final analytic sample, which are a subsample of the previous study. Bottom row: Distress trajectories within the DTI subsample (N=7,581). Left-most panel shows all trajectory classes; remaining panels show each trajectory class separately. Thick/bolded lines indicate mean score within each trajectory; faded lines indicate individual participant scores. Visits indicate annual study visits. For visualization purposes, Distress scores greater than 80 are not shown (removal of N=7 data points).

Figure S3. Associations between all T1-weighted and DTI Psychosis Neuroscores and Distress PLE trajectory membership.

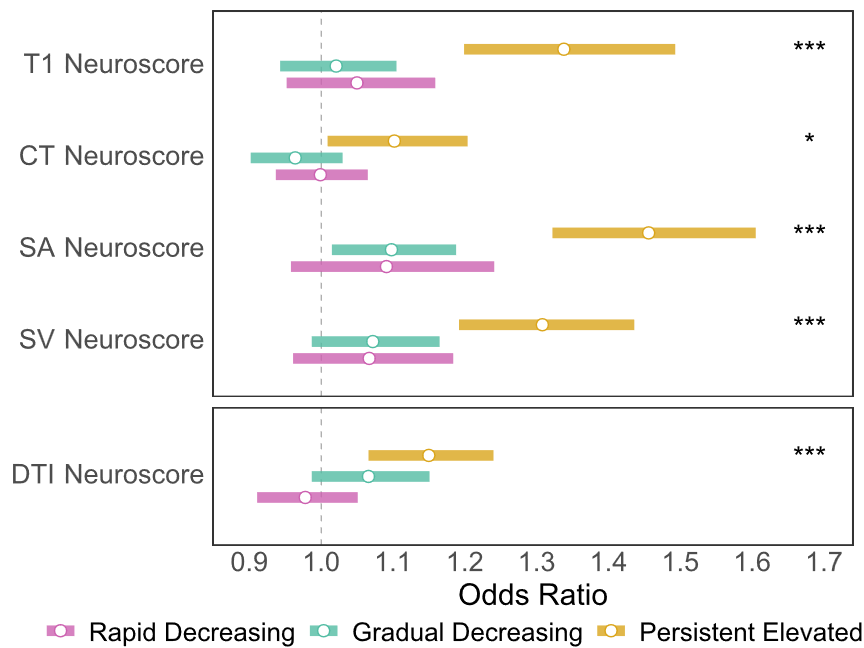

Reference trajectory is the Low/No Distress trajectory. T1 Neuroscore is the sum of cortical thickness, surface area and subcortical volume measures; DTI Neuroscore comprises fractional anisotropy measures. Circles represent odds ratios; bars represent 95% confidence intervals. Asterisks indicate significant difference (FDR-corrected) between corresponding trajectory and reference trajectory (the Low/No Distress trajectory). Abbreviations: CT, cortical thickness; CI, confidence interval; *d*, Cohen’s *d*; DTI, diffusion tensor imaging; OR, odds ratio; SA, surface area; SV, subcortical volume. * = *p*_FDR_<0.05; ** = *p*_FDR_<0.01; *** *p*_FDR_<0.001.

Figure S4. Associations between T1-weighted and DTI Psychosis Neuroscores and Distress PLE trajectory membership, after mutually adjusting for each Neuroscore.

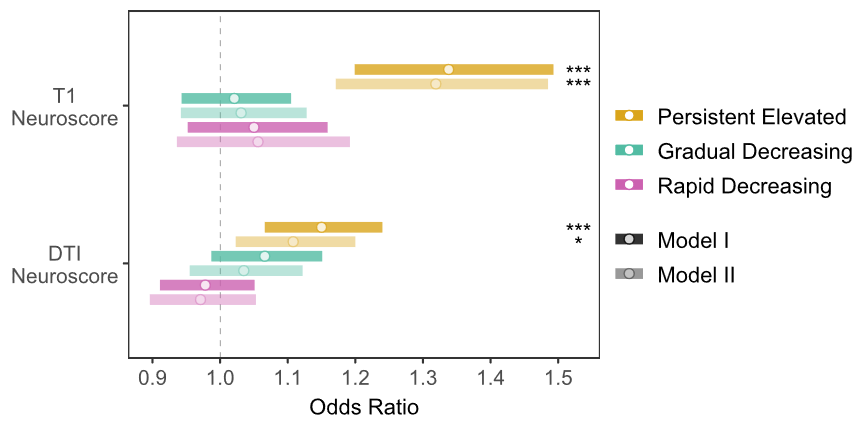

Odds ratios for associations between T1-weighted and DTI Neuroscores and Distress PLE trajectory membership. Model I indicates original model; Model II adjusted for respective T1/DTI Psychosis Neuroscore in same model (i.e., T1 Neuroscore adjusted for DTI Neuroscore in same model, and vice versa). Circles represent odds ratios; bars represent 95% confidence intervals. Reference trajectory is the Low/No Distress trajectory. * = *p*_FDR_<0.05; ** = *p*_FDR_<0.01; *** *p*_FDR_<0.001.

Figure S5. Associations between modifiable risk factors and Psychosis Neuroscores and Distress PLE trajectory membership.

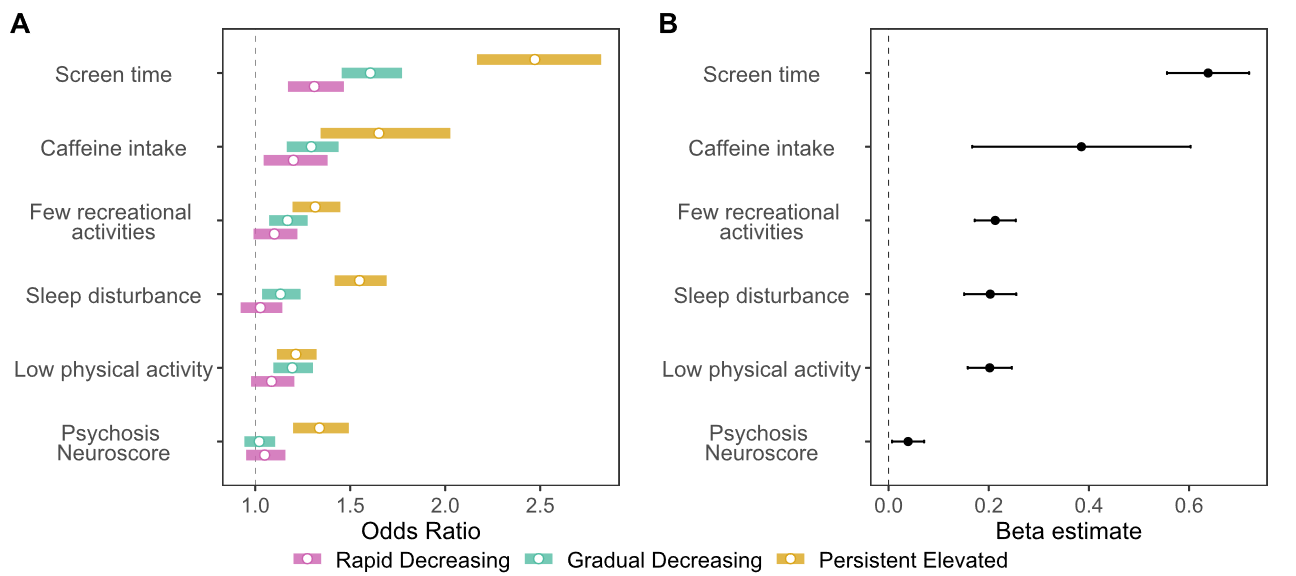

**A.** Odds ratios for separate models examining associations between modifiable risk factors and T1-weighted Psychosis Neuroscores and Distress PLE trajectory membership. Circles represent odds ratios; bars represent 95% confidence intervals. Reference trajectory is the Low/No Distress trajectory**. B.** Model coefficients from a multivariable model examining associations between modifiable risk factors and the T1-weighted Psychosis Neuroscore in predicting trajectory group membership (as a latent variable). Points indicate beta coefficients; bars indicate standard errors.
